# Drivers of Diagnostic Delay in Mitochondrial Disease: Missed Recognition of Canonical Features

**DOI:** 10.1101/2025.10.09.25337582

**Authors:** Rory J Tinker, Neil Jacob, Mohammad Ghouse Syed, Janhawi Kelkar, Colleen Donnelly, Ibrahim Elsharkawi, Jaya Ganesh, Bruce Gelb, Vikas Pejaver, Tamas Kozicz, Eva Morava

**Author notes:** Equal contributions (Rory J Tinker, Neil Jacob). CORRESPONDING AUTHOR: **Eva Morava,**.

## Abstract

**Background:** Diagnostic delay is common in mitochondrial disease, and its drivers remain unclear despite advances in molecular diagnostics.

**Methods:** We retrospectively analyzed 58 individuals with molecularly confirmed mitochondrial disease at the Mount Sinai Mitochondrial Disease Clinic, diagnosed after 2016. Diagnostic delay was partitioned into intervals from symptom onset to clinical suspicion, and from suspicion to molecular diagnosis. Demographic, phenotypic, and genetic data were abstracted from health records, and Human Phenotype Ontology terms were compared before and after diagnosis using ClinPhen.

**Results:** Most delays occurred between symptom onset and clinical suspicion (mean 8.17 years) rather than after suspicion (mean 0.63 years), yielding a mean total delay of 8.8 years (median 3.0). Delay decreased sharply by year of birth (r = –0.99, p < 1.5 × 10^–48) and symptom onset (r = –0.98, p < 1.4 × 10^–36), but showed no meaningful trend with year of diagnosis. Developmental delay predicted shorter diagnostic intervals. Canonical features such as seizures, hypotonia, and stroke were frequently documented years before suspicion, underscoring missed opportunities.

**Conclusions:** Diagnostic delay reflects missed recognition rather than testing limitations. Systematic capture of early phenotypes and AI/NLP-based mining of electronic health records could proactively flag patients for reflexive sequencing, shortening diagnostic delay.

## Introduction

Primary mitochondrial diseases are a clinically and genetically heterogeneous group of disorders affecting cellular energy metabolism by pathogenic variants in mitochondrial DNA (mtDNA) or nuclear genes. ^1–3^ They can involve virtually any organ system, usually with multi-organ manifestations, although isolated organ involvement can occur. Organ systems most commonly affected include the central nervous system, skeletal muscle, ^4^ Clinical manifestations and age of onset are highly variable,which may delay uniform recognition of these disorders. ^5^ Diagnostic delays arise because early features are nonspecific, the course of disease is variable, and due to a lack of robust diagnostic biomarkers. ^6^ Early and accurate diagnosis is essential for patient management, reproductive counseling, and access to emerging therapies, yet mitochondrial disease remains under-recognized across both pediatric and adult populations. ^2^.

Most existing studies on diagnostic delay in mitochondrial disease originate from single centers or focus on specific syndromic subtypes, often lacking standardized definitions of the diagnostic interval or systematic assessment of associated clinical and demographic factors. ^7–11^ A recent scoping review of monogenic disease highlighted substantial variability in study design, outcome measures, and conclusions, revealing that inconsistent definitions of delay severely limit cross-study comparability and broader. ^12^ In parallel, a novel conceptual model applied to Mendelian disease within electronic health records now defines discrete diagnostic intervals (pre-suspicion, pre-diagnosis, and post-diagnosis) as captured by the EHR, enabling systematic study of diagnostic delay in mitochondrial disease. ^13^

In this pilot study, we examined diagnostic delay in a clinically and genetically diverse cohort of individuals with clinically confirmed mitochondrial disease, using the electronic health record (EHR) to reconstruct the diagnostic trajectory. Our primary outcome was the interval from symptom onset to genetic diagnosis. To better resolve this trajectory, we further partitioned the overall delay into two sequential phases, following a recently proposed conceptual model: (1) the interval from symptom onset to first clinical suspicion of mitochondrial disease and (2) the interval from clinical suspicion to confirmed genetic diagnosis. We examined how these intervals varied by age group, sex, molecular etiology (mtDNA versus nuclear), and clinical phenotype. In addition, we applied natural language processing (NLP) to extract Human Phenotype Ontology (HPO) terms from electronic health records, enabling systematic comparison of phenotype prevalence before and after clinical suspicion for mitochondrial disease. We found that most delay arose before a specific clinical suspicion for a possible mitochondrial diagnosis, that developmental delay was linked to shorter diagnostic intervals, and that canonical features such as seizures, hypotonia, and stroke were often present years before clinical suspicion and diagnosis.

## Methods

### Cohort construction

This study included individuals evaluated at Mitochondrial Medicine Program at Mount Siani Hospital. Only patients diagnosed between 2016 and 2025 were included (IRB 25-00728 Icahn School of Medicine at Mount Sinai Institutional Review Board.). ^14^ Eligible participants had a confirmed clinical diagnosis of primary mitochondrial disorders as determined by a board-certified clinical biochemical geneticist. We exclude individuals with secondary mitochondrial dysfunction due to another primary genetic diagnosis or an acquired condition. Clinical and genetic data were abstracted from the electronic medical record and supplemented by review of gene testing reports. When available, we also collected demographic, clinical, and laboratory variables for exploratory analyses, including sex, genotype category (nuclear vs mitochondrial), clinical diagnosis, number of similarly affected family members, developmental delay, seizure history, and metabolic abnormalities including elevated lactate. All data were de-identified before analysis.

### Definition of diagnostic delay and clinical suspicion

Diagnostic delay and date of clinical suspension were defined and dated based on a previous model. ^13^ Diagnostic delay was the time from reported symptom onset to the age at confirmed molecular diagnosis of mitochondrial disease. Symptom onset was abstracted from the medical record using patient or caregiver reports and clinician assessment of when relevant features first appeared. Clinical diagnosis was the date a pathogenic or likely pathogenic variant was identified by genetic testing and confirmed by a clinical genetics’ provider. Clinical suspicion was the earliest date a clinician documented that a primary mitochondrial disorder or another genetic disorder was being actively considered and warranted evaluation. We identified this date from signed notes or the problem list using explicit phrases such as “suspect mitochondrial disease,” “genetic etiology under consideration,” or a named syndrome, and from plans that initiated targeted evaluation for mitochondrial or genetic disease. When explicit language was absent, we used the earliest strong proxy indicating intent to evaluate for a mitochondrial or genetic disorder, including referral to a mitochondrial clinic or clinical genetics service, ordering mtDNA or nuclear panels or exome or genome sequencing, or starting a diagnostic workup directed at mitochondrial or genetic disease. Dates came from the note timestamp for documentation, the order date for tests, and the referral date for specialty evaluations.

### Phenotypic Data Extraction

Clinical notes for each individual were retrieved from the Mount Sinai Health System using the AI-Ready Mount Sinai (AIR.MS) platform (https://labs.icahn.mssm.edu/airms/). The corpus included progress notes, care notes, telephone encounters, and other document types. Phenotypic features were extracted with ClinPhen, a natural language processing tool that maps clinical text to Human Phenotype Ontology (HPO) terms

### Statistical Analysis

We performed a cross-sectional analysis of diagnostic delay. Descriptive statistics were calculated for the overall cohort, including means, standard deviations (SD), standard errors of the mean (SEM), and medians for continuous variables, and frequencies and percentages for categorical variables. To evaluate factors associated with diagnostic delay, variables were categorized into three groups: (1) continuous clinical and demographic variables, (2) binary clinical features (e.g., presence or absence of developmental delay or seizures), and (3) categorical variables with more than two levels (e.g., clinical diagnosis, genetic etiology). Pearson correlation coefficients were used to assess associations between continuous variables and diagnostic delay. For binary and categorical variables, independent-sample t-tests and one-way analysis of variance (ANOVA) were used to compare mean diagnostic delay between groups. To characterize missed diagnostic opportunities, HPO terms were analyzed for frequency and mean lead time (years before suspicion or diagnosis), and their prevalence before and after suspicion was compared. Results were visualized with dumbbell plots and summarized in supplementary tables. All analyses were conducted in R (version 4.4.1), with results exported into structured Excel files for integration into figures and manuscript tables. ^15^ For each patient, only the first occurrence of an HPO term was retained for downstream analyses (**Figure 1**). ClinPhen is openly available (https://github.com/kingmanzhang/clinphen/tree/master) and was run in Python (version 3.11.11). Among the 58 genetically confirmed individuals, clinical note data were available for 50. A total of 20,702 notes were retrieved from 49 individuals, while one individual had no available notes. After restricting to the first occurrence of each term per individual, the dataset comprised 5,751 HPO terms across all patients, representing 1,489 unique terms.

**Figure 1.**
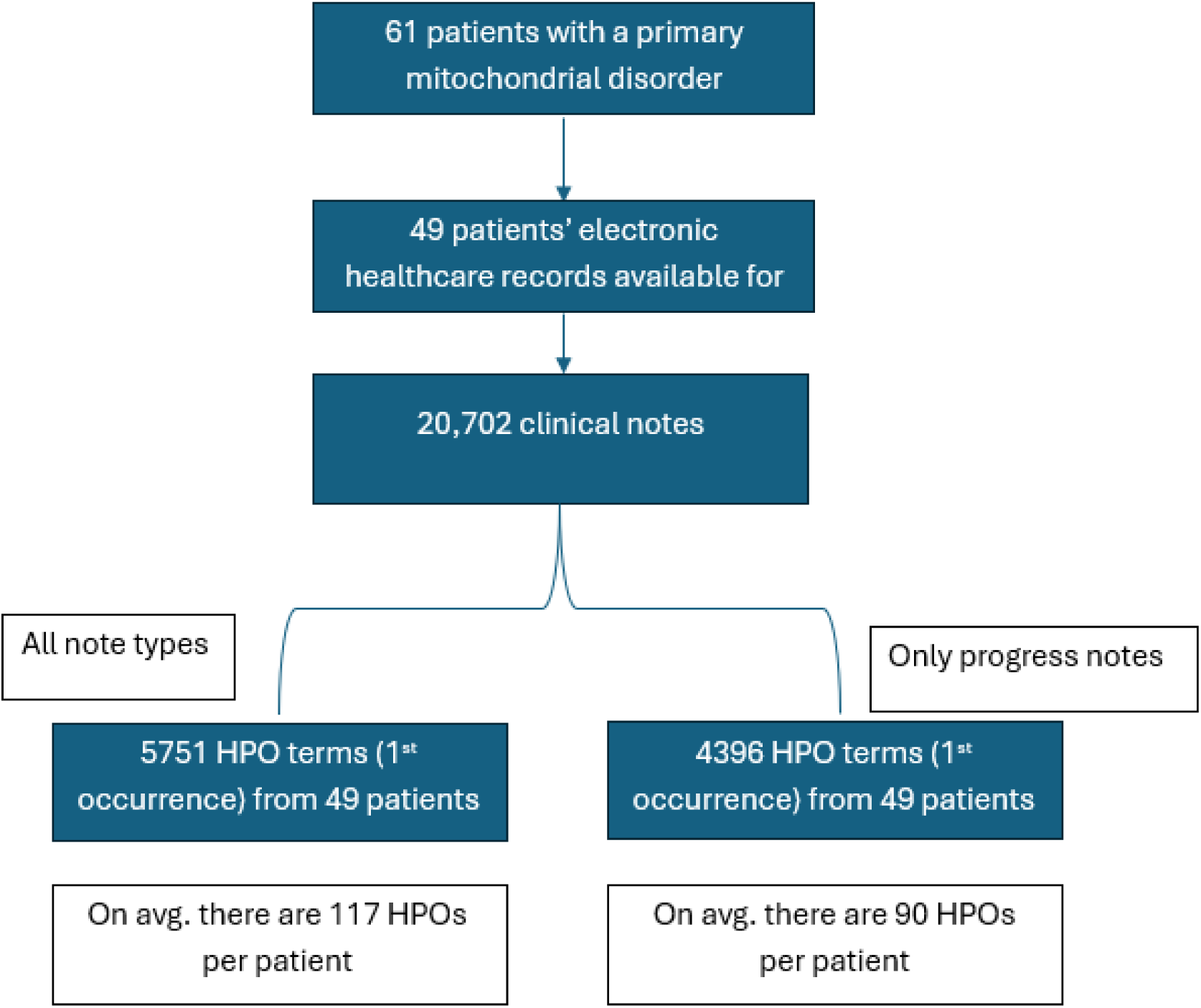
Cohort flow chart and Human Phenotype Ontology (HPO) term extraction. Flow diagram showing cohort selection and phenotypic feature extraction. Of 61 individuals with a clinically confirmed primary mitochondrial disorder, electronic health records were available for 49 patients, yielding 20,702 clinical notes. Using ClinPhen, a total of 5,751 first-occurrence HPO terms were extracted across all note types (mean 117 HPO terms per patient). Restricting analysis to progress notes only resulted in 4,396 first-occurrence HPO terms (mean 90 HPO terms per patient).

## Results

### Clinical and Genetic Features of the Cohort

This study included 58 individuals with molecularly confirmed mitochondrial disease evaluated at Mitochondrial Medicine Program at Mount Sinai Hospital. The age range and mean age at chart review were 0.5-77 years and 22.5 years, respectively. Two individuals were deceased and their ages at death were used. The age of symptom onset ranged from 0 – 42 years old with a mean age of 9.7 years. The age of symptom onset was unknown in two individuals and age of genetic diagnosis was unknown in one individual. The mean age at the time of genetic diagnosis was 18.5 years (n = 57) (**Figure 1**). The cohort was approximately evenly split by sex, with 31 females (53%) and 27 males (47%). Genetic etiologies were nearly evenly distributed between mitochondrial DNA–based disorders (62%, n=36) and nuclear gene–based disorders (38%, n=22). Heteroplasmy in individuals affected by mitochondrial DNA-based disorders ranged from 5-100%.

Mitochondrial DNA–based disorders, which accounted for 62% (n = 36) of cases, were divided into disorders caused by point mutations (94%, n = 34/36) or by deletions (6%, n = 2/36). The **m.3243A>G** variant, classically associated with *mitochondrial encephalomyopathy, lactic acidosis, and stroke-like episodes* (**MELAS**), was the most common (67%, n = 23/34). The second most common variant was **m.8363G>A** (11.7%, n = 4/34), typically seen in *myoclonic epilepsy with ragged-red fibers* (**MERRF**). Other point mutation included m.3302A>G (MT-TK, n=1/34), m.9997T>A (MT-TG, n=1/34), m.3460G>A (MT-ND1, n=1/34), m.11778G>A (MT-ND4, n=1/34), m.14484T>C (MT-ND6, n=1/34), m.3302A>G (tRNA[Leu(UUR)], n=1/34), m.8993T>G (MT-ATP6, n=1/34), and m.8719G>A (MT-ATP, n=1/34) (**Table 1**). Of the individuals with a point mutation, 17.6% (n=6/34) were asymptomatic, but were identified with the genetic variant after familial cascade testing. This included 4 individual families. One individual was incidentally identified with the m.14484T>C variant after genetic testing for what is believed to be unrelated congenital arthrogryposis.

**Table 1.** Cohort Characteristics and Clinical Subgroups.

| **Table 1. Cohort Characteristics and Clinical Subgroups** |  |
| --- | --- |
| **Characteristics** | **Value** |
| Total Participants | 58 |
| Age Range | 0.5 - 77 |
| Mean Age (years) (2 individuals were deceased, their age at death were used) | 22.5 |
| Age at onset, range (years) | 0 – 42 |
| Mean Age of Onset (years) | 9.7 (2 unknown) |
| Mean Age at Genetic Diagnosis | 18.5 (1 unknown) |
| Interval: Onset --> Clinical Suspicion (years) | 8.17 (2 unknown) |
| Females | 31 (53%) |
| Males | 27 (47%) |
| **Mitochondrial** | **36 (62%)** |
| Mt Deletion | 2 |
| Mt Point Mutations | 34 |
| m.3243A>G | 23 |
| m.8363G>A | 4 |
| m.9997T>A | 1 |
| m.11778G>A | 1 |
| m.3302A>G | 1 |
| m.8993T>G | 1 |
| m.8719G>A | 1 |
| m.3460G>A | 1 |
| m.14484T>C | 1 |
| **Nuclear** | **22 (38%)** |
| Transport Protein | 2 |
| *SLC25A4* | 2 |
| Small Molecule Metabolism | 6 |
| *ECHS1* | 3 |
| *PC* | 1 |
| *DLD* | 1 |
| *PPA2* | 1 |
| Complex I | 3 |
| *NDUFV1* | 2 |
| *NDUFAF1* | 1 |
| Complex IV | 1 |
| *SURF1* | 1 |
| Complex V | 1 |
| *TMEM70* | 1 |
| Translation Defects | 6 |
| *GTPBP3* | 1 |
| *MTO1* | 1 |
| *NARS2* | 1 |
| *TRIT1* | 1 |
| Mitochondrial Structure/Integrity | 3 |
| *OPA1* | 1 |
| *TAZ* | 1 |
| Mitochondrial Maintenance/Integrity | 3 |
| *POLG* | 2 |
| *TOP3A* | 1 |

Nuclear-encoded disorders (38%, n=22) were subcategorized into defects in protein transport (*SLC25A4*), small molecular metabolism (*ECHS1,* Pyruvate Carboxylase Deficiency, Dihydrolipoamide Dehydrogenase Deficiency, and Inorganic Pyrophosphatase Deficiency), protein translation *(GTPBP3, TRIT1, MTO1, NARS2),* mitochondrial structure/integrity *(OPA1*, *TAZ*), mitochondrial maintenance/integrity (*POLG, TOP3A),* and nuclear complex function (*NDUFAF1, NDUFV1, SURF1, TMEM70)* (**Table 1**).

### Diagnostic Delay Predominantly Reflects Lag in Clinical Suspicion

The mean delay was 8.8 years (SD: 9.9, SEM: 1.29; n = 58), highlighting the prolonged but variable time to diagnosis experienced by many individuals with mitochondrial disease (**FIGURE 2**). The mean age of symptom onset was 9.7 years (n=58) with clinical suspicion typically raised at a mean age of 17.0 years (n = 61). On average, the interval from first symptom onset to clinical suspicion was 8.17 years of the delay of, while the interval from clinical suspicion to molecular diagnosis was 0.63 years. These diagnostic timing data are summarized in Table 1 and graphically represented in **Figure 2**.

**Figure 2.**
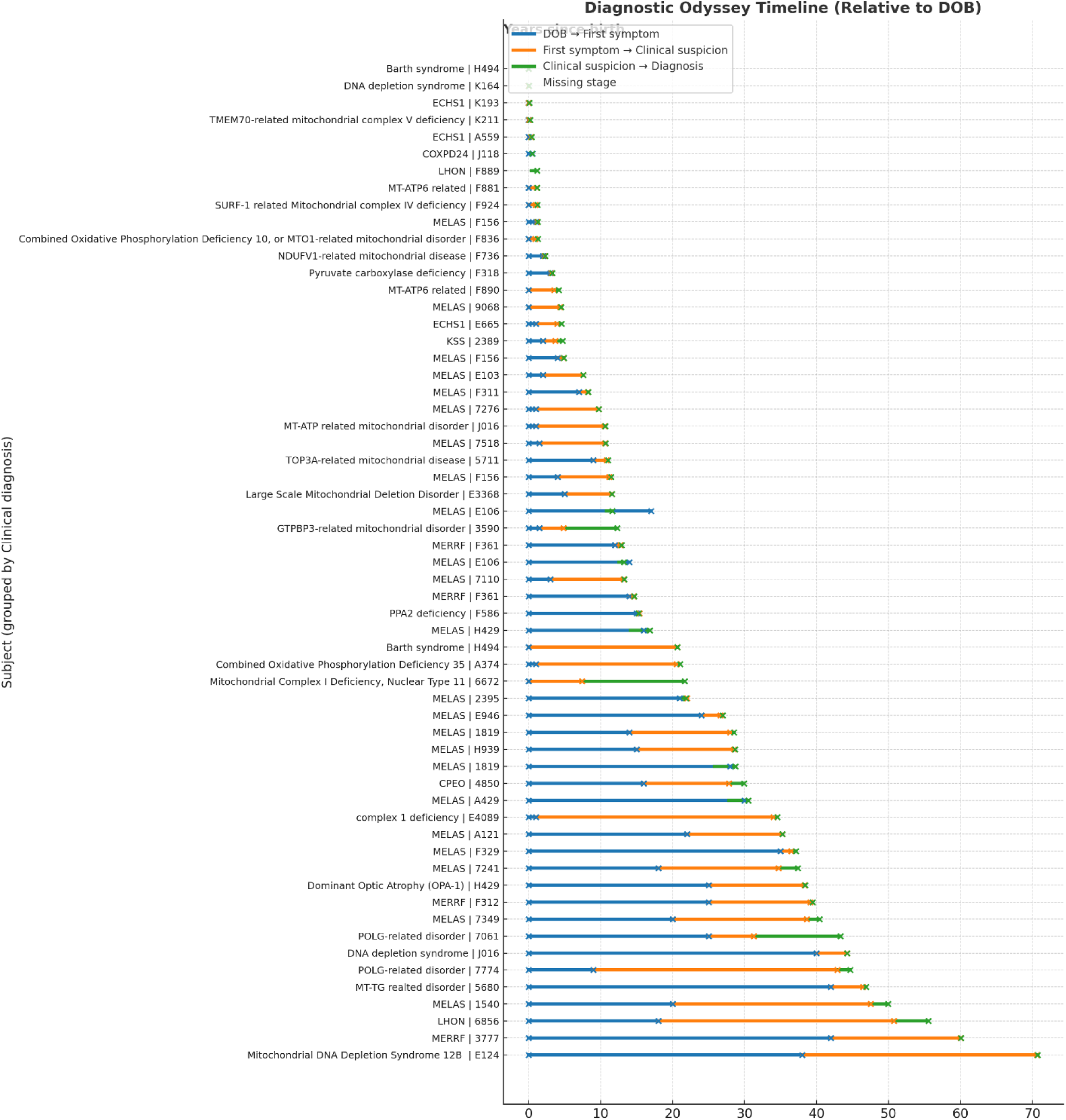
Diagnostic odyssey timeline of individuals with mitochondrial disease. Each horizontal bar represents a single participant, grouped by clinical diagnosis, showing the timeline from birth to key diagnostic milestones. Blue segments represent the time from birth to first reported symptom onset, orange segments represent the interval from symptom onset to first clinical suspicion, and green segments represent the interval from clinical suspicion to molecular diagnosis. Circles mark the specific ages at which each stage occurred. Missing stages are indicated by gaps in the timeline. The figure highlights the substantial variability in diagnostic trajectories across individuals, with most of the delay arising during the interval from symptom onset to clinical suspicion.

### Birth and Onset Year Predict Shorter Delays, While Diagnosis Year Does Not

Diagnostic delay has shortened substantially across successive generations of patients with mitochondrial disease. As shown in **Figure 3A**, there was a strong inverse correlation between year of birth and diagnostic delay (r = –0.99, p < 1.5 × 10^–48), with later-born individuals experiencing markedly shorter delays. This pattern was confirmed when the cohort was split by birth cohort halves, with those born earlier experiencing significantly longer delays compared to those born more recently (**Figure 3B**, p = 7.3 × 10^–12). In contrast, the year of molecular diagnosis was not associated with diagnostic delay (**Figure 3C**, r = 0.03, p = 0.84), and splitting the cohort by earlier versus later diagnosis years did not reveal a difference (**Figure 3D**, p = 0.98). Instead, the key driver of improvement was the era of symptom onset: diagnostic delay was strongly negatively correlated with year of onset (**Figure 3E**, r = –0.98, p < 1.4 × 10^–36), and individuals with later-onset symptoms had significantly shorter delays compared to those with earlier onset (**Figure 3F**, p = 1.3 × 10^–8). Collectively, these findings indicate that diagnostic delays have decreased over time, largely due to changes in when symptoms arise and are first recognized, rather than improvements tied directly to the calendar year of diagnosis.

**Figure 3.**
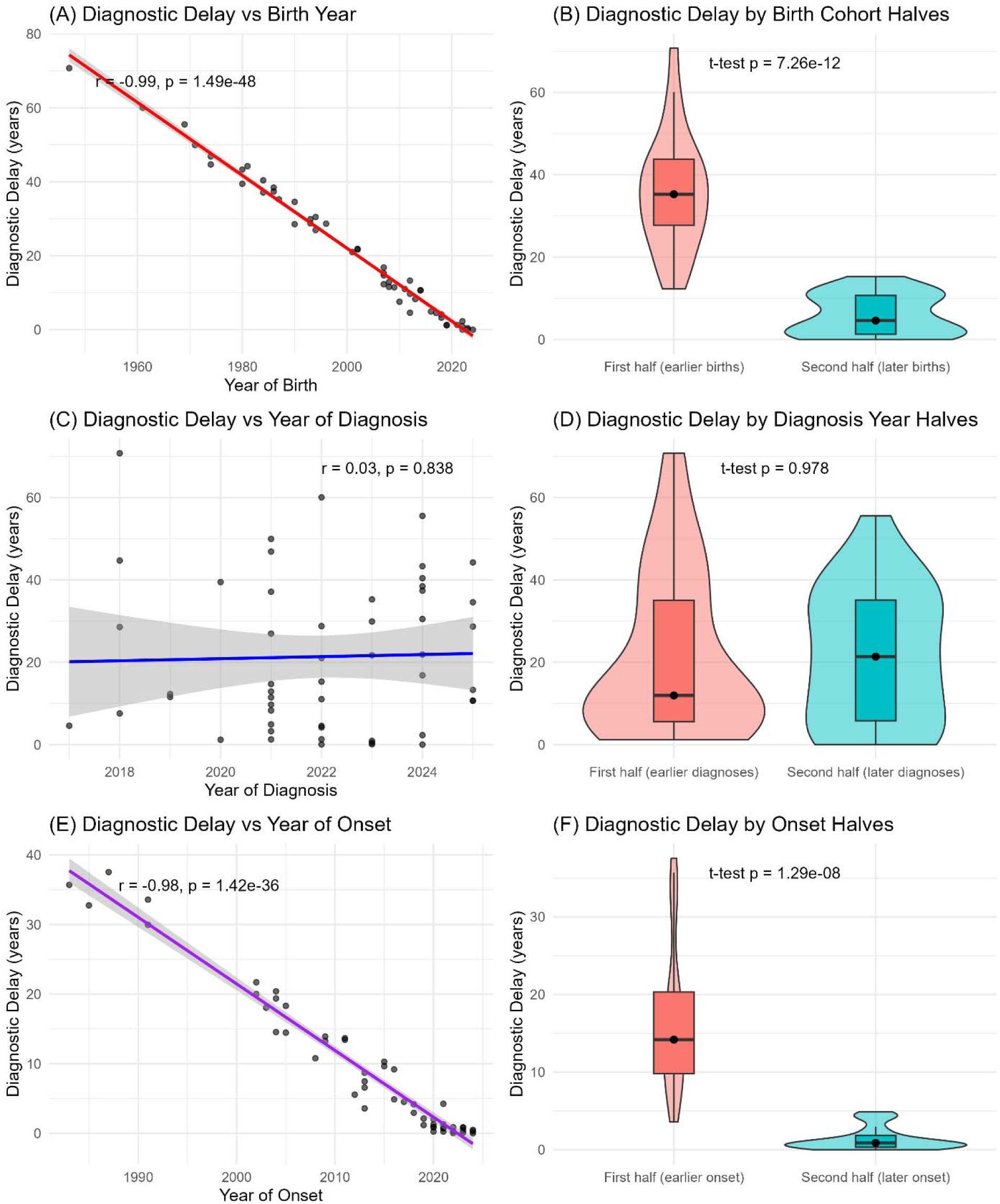
Trends in diagnostic delay across birth year, diagnosis year, and symptom onset. (A) Diagnostic delay was strongly negatively correlated with year of birth (r = –0.99, p < 1.5 × 10^–48), with later birth cohorts experiencing shorter delays. (B) Patients born in the later half of the cohort had significantly shorter delays compared with those born earlier (p = 7.3 × 10^–12). (C) Diagnostic delay showed no significant association with year of molecular diagnosis (r = 0.03, p = 0.84). (D) Splitting by earlier versus later diagnosis year revealed no difference in delay (p = 0.98). (E) Diagnostic delay was strongly negatively correlated with year of symptom onset (r = –0.98, p < 1.4 × 10^–36). (F) Patients with later symptom onset experienced shorter delays compared with those with earlier onset (p = 1.3 × 10^–8). Together, these panels show that diagnostic delays have decreased substantially over time, primarily due to differences in birth cohort and age of symptom onset, rather than year of diagnosis.

### Factors positively associated with increased diagnostic delay

We next examined demographic and clinical factors that might predict variation in diagnostic delay (**Figure 4**). Several features emerged as significant predictors. Brainstem involvement was associated with markedly shorter diagnostic delays (mean difference 8.5 years, p = 1.2 × 10^–6), and developmental delay at presentation was likewise linked to shorter delays (mean difference 11.1 years, p = 0.019). In contrast, visual loss was associated with significantly longer delays (mean difference 5.9 years, p = 0.011). Finally, later age of symptom onset correlated with longer diagnostic delays (r = 0.29, p = 0.033), suggesting that mitochondrial disease presenting in older individuals is more easily overlooked. By contrast, neither sex nor the broad category of nuclear versus mitochondrial DNA genetic etiology was associated with time to diagnosis. Several additional variables were tested but were not significant predictors, including common clinical features (such as seizures, cardiomyopathy, and muscle weakness), laboratory parameters, and age at first presentation. These non-significant associations are shown in **Supplementary Figure 1** and **Supplementary Table 1.**

**Figure 4.**
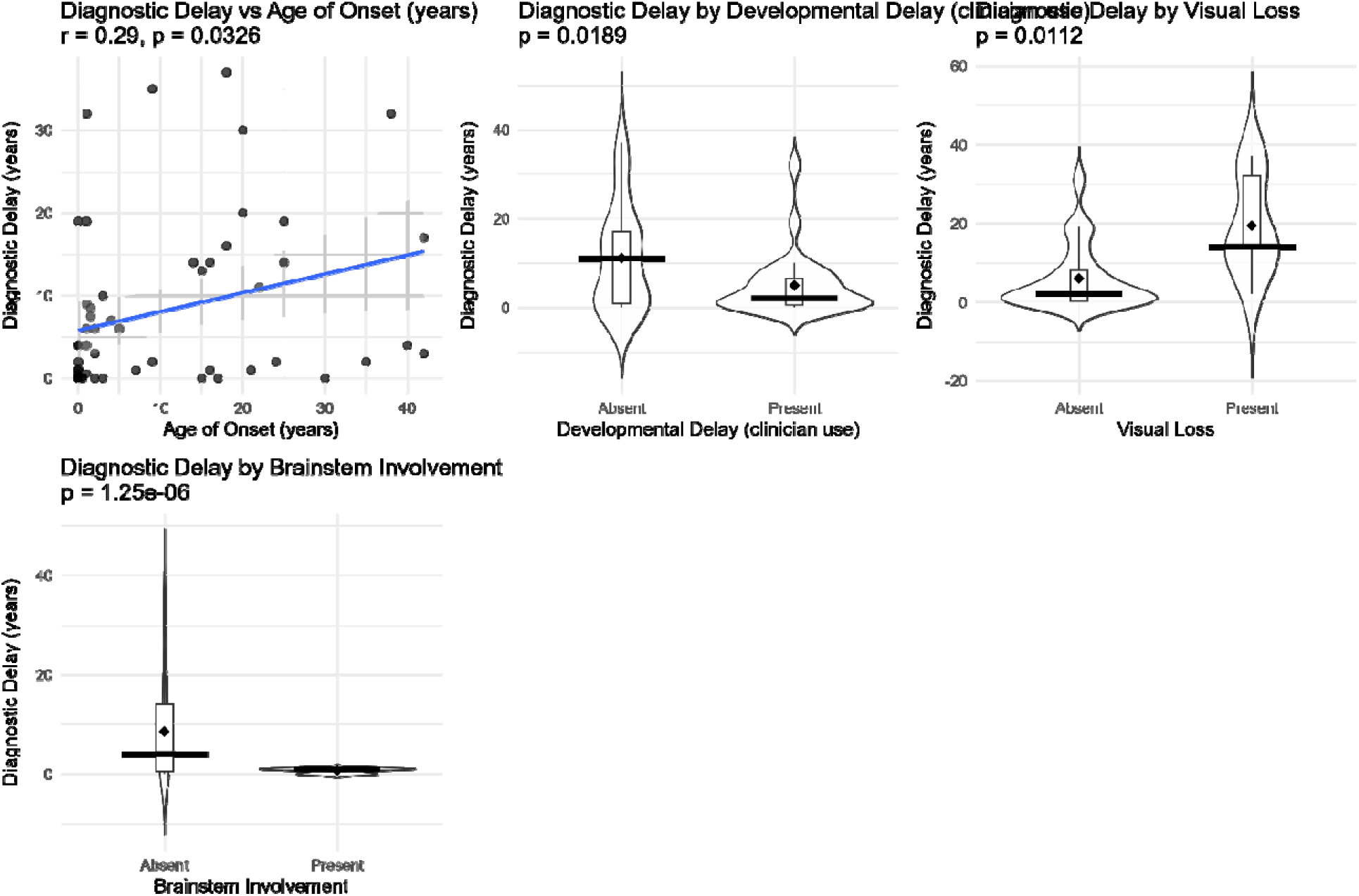
Significant clinical and demographic features associated with diagnostic delay. Plots showing features significantly associated with diagnostic delay in mitochondrial disease. **Brainstem involvement** and **developmental delay** were associated with shorter diagnostic delays, while **visual loss** was associated with shorter delays. **Younger age of onset** correlated with shorter delay, whereas older onset correlated with longer delay. Scatter, violin, and box plots illustrate distributions with mean and median estimates, and p-values are from t-tests or Pearson correlation as appropriate.

### Sub-analysis of Familial Cases

Of the 58 cases, five were identified through familial cascade testing (**Supplement table 2**), including three with MELAS and two with MERRF. The mean age at diagnosis in these cases was 14.4 years, earlier than in non-familial cases though not statistically significant (18.5 years; *p*<0.40, unpaired *t* test). The chart review showed that two of the five patients had clinical symptoms prior to diagnosis, while three developed symptoms after their genetic diagnosis.

### Documented HPO Features Reveal Missed Diagnostic Opportunities

Analysis of clinical documentation revealed that several key HPO terms were frequently present in the medical record well before mitochondrial disease was suspected and diagnosed (**Figure 5, Supplementary Table 3**). Canonical features such as seizures (mean 1.8 years before diagnosis; 41% of individuals), global developmental delay (3.7 years; 36%), hypotonia (2.1 years; 36%), and stroke (4.0 years; 27%) were consistently documented years in advance, highlighting a substantial diagnostic window. Other features, including neonatal hypotonia (4.6 years; 23%), hearing impairment (0.6 years; 18%), vomiting (1.9 years; 23%), and increased body weight (1.7 years; 27%), were also recorded prior to suspicion and often persisted. By contrast, nonspecific symptoms such as fatigue, constipation, cough, anxiety, and depression were common but less clearly disease-related, with shorter lead times and limited specificity. Together, these findings demonstrate that diagnostically informative phenotypes were often present in the EHR long before recognition, suggesting that systematic capture and flagging of such terms could substantially shorten diagnostic delays.

**Figure 5.**
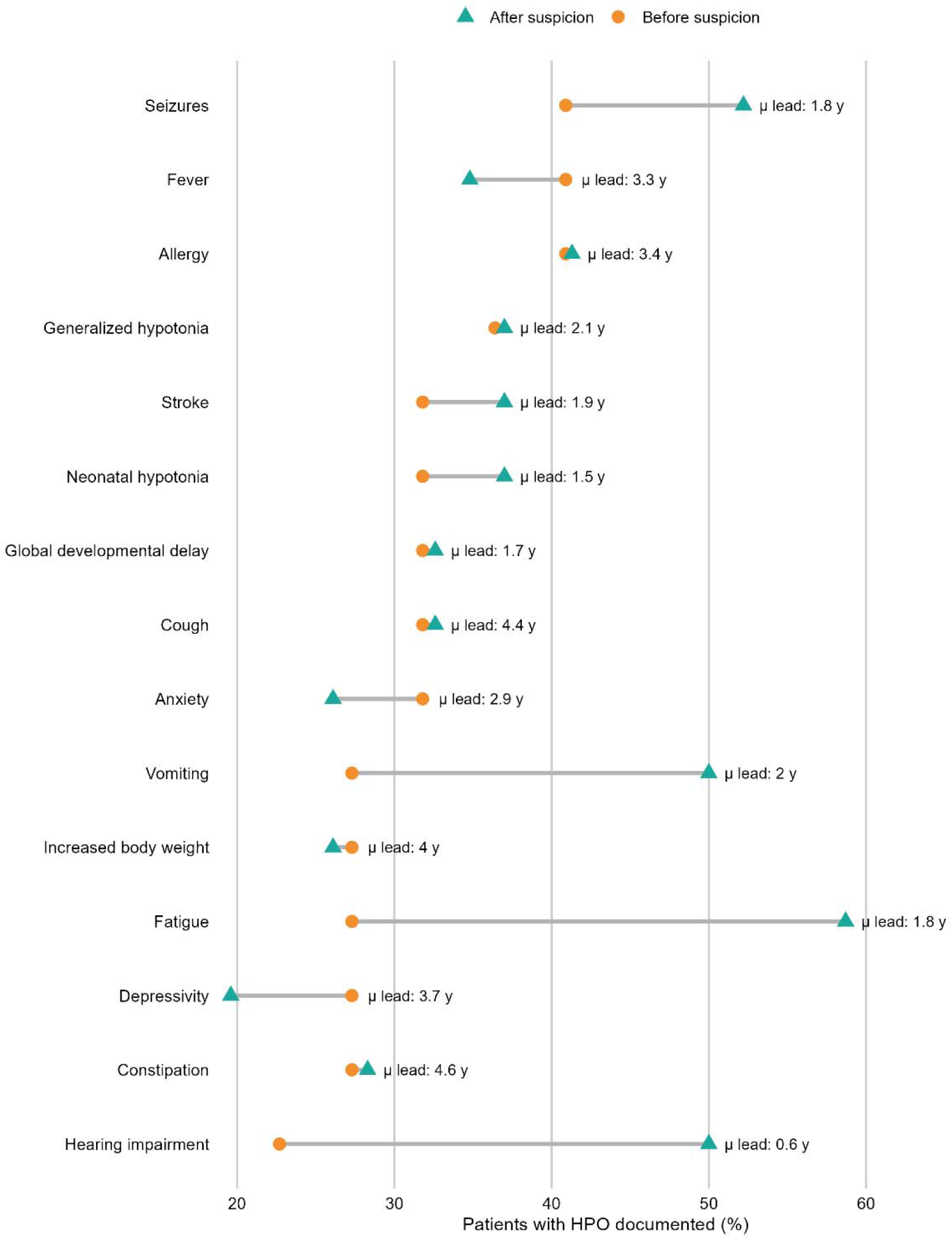
HPO features documented before vs. after clinical suspicion of mitochondrial disease. Dumbbell plot showing the proportion of patients with selected Human Phenotype Ontology (HPO) terms documented before (circle, left) and after (triangle, right) the point of clinical suspicion for mitochondrial disease. Features such as seizures, global developmental delay, hypotonia, and stroke were frequently recorded years in advance, while nonspecific features (e.g., fatigue, constipation, anxiety) were also common but less clearly interpreted as disease-related. Horizontal connectors illustrate persistence of symptoms across both periods, highlighting missed opportunities for earlier recognition. Mean lead time (years prior to diagnosis) is displayed adjacent to each feature.

Demographic, genetic, and diagnostic characteristics of the 61 individuals with molecularly confirmed mitochondrial disease evaluated at the Mount Sinai Mitochondrial Disease Clinic. Continuous variables are summarized as mean ± standard deviation (SD). Intervals represent elapsed time between symptom onset, first clinical suspicion, and molecular genetic diagnosis. Sex distribution, genetic etiology (mitochondrial vs nuclear), and major clinical subgroups are listed with corresponding sample sizes.

## Data Availability

Deidentified data available upon reasonable request.

## Abbreviations

(AI): Artificial intelligence
(EHR): electronic health record
(GINA): Genetic Information Nondiscrimination Act
(HPO): Human Phenotype Ontology
(ML): machine learning
(NLP): natural language processing
(mtDNA): mitochondrial DNA
(WES): wholeexome sequencing
(WGS): whole-genome sequencing
(MELAS): mitochondrial encephalomyopathy, lactic acidosis, and stroke-like episodes

## Acknowledgements

M. G. S. and V. P. were supported by NIH/NLM grant R00LM012992. This work was supported in part through the Minerva computational and data resources and staff expertise provided by Scientific Computing and Data at the Icahn School of Medicine at Mount Sinai and supported by the Clinical and Translational Science Awards (CTSA) grant UL1TR004419 from the National Center for Advancing Translational Sciences. Research reported in this publication was also supported by the Office of Research Infrastructure of the National Institutes of Health under award number S10OD026880 and S10OD030463. The content is solely the responsibility of the authors and does not necessarily represent the official views of the National Institutes of Health.

## Data availability

De-identified patient-level data underlying the findings of this study are available from the corresponding author upon reasonable request, subject to institutional approvals and compliance with applicable regulations to protect patient privacy.

## Conflict of Interest Statement

The authors declare that they have no relevant conflicts of interest related to this work.

## Supplementary Figures and table Legends

**Supplementary Figure 1a–b.**
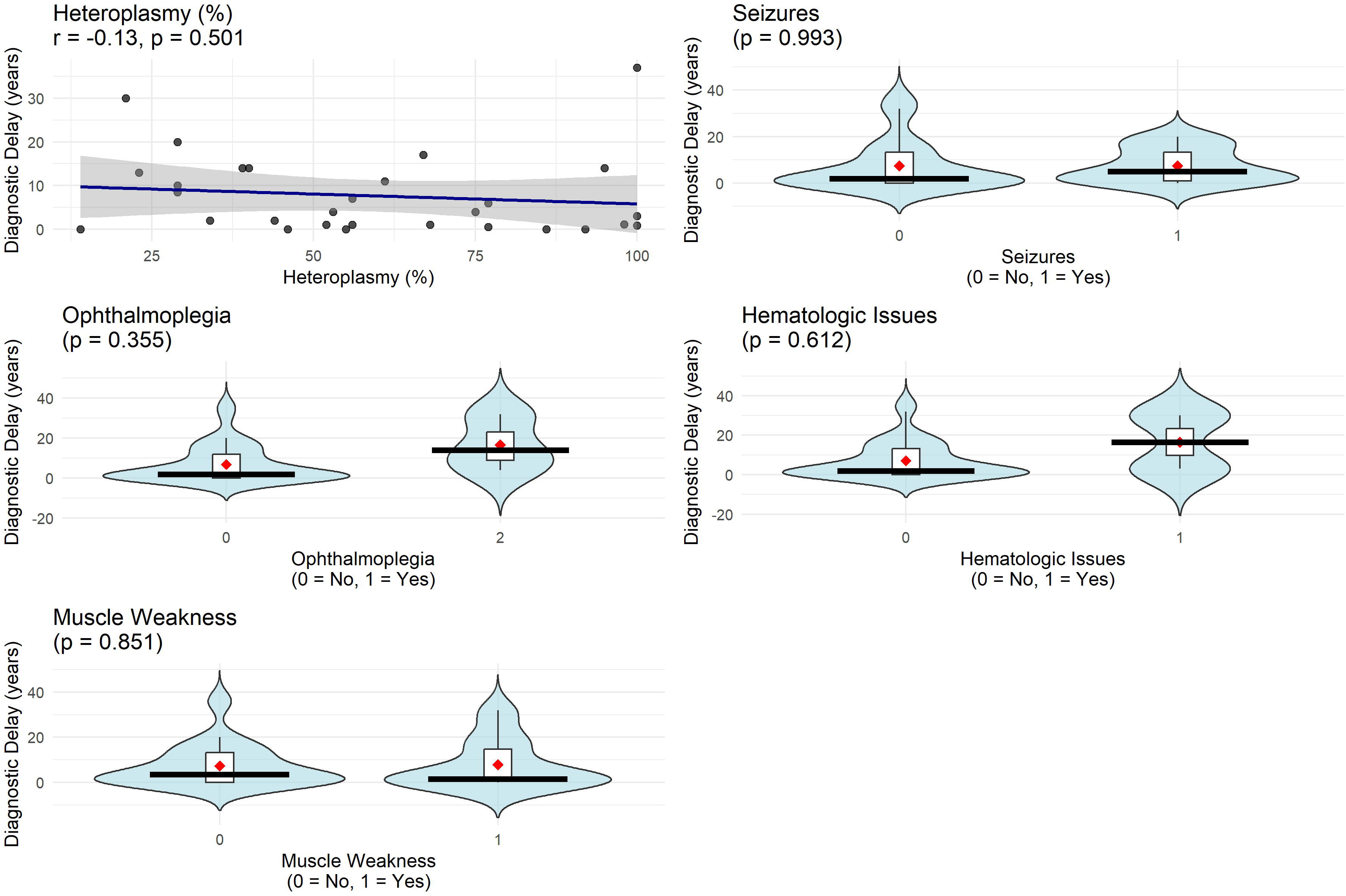
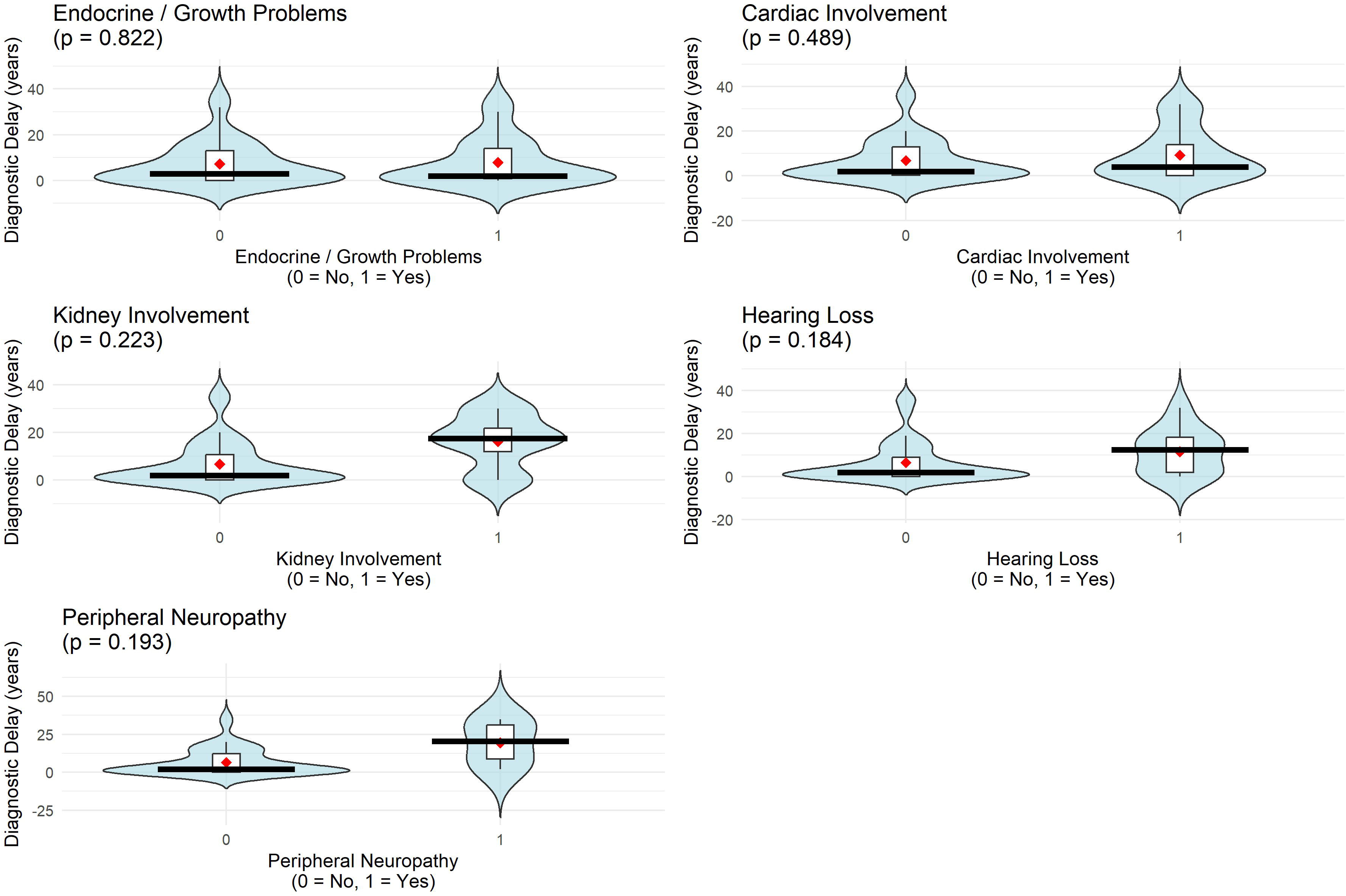
Expanded visualizations of additional clinical features that were not significantly associated with diagnostic delay. Binary features are displayed as violin plots comparing individuals without (0) and with (1) the feature; the red diamond marks the group mean, and boxplots indicate the median and interquartile range. Heteroplasmy (%) is shown as a scatterplot with fitted regression line and 95% confidence band. None of the associations reached statistical significance (p < 0.05).

**Supplementary Table 1. Predictors of diagnostic delay in mitochondrial disease.** Summary of demographic, genetic, and clinical features tested for association with diagnostic delay. The table reports sample size, test statistics, and p-values for each factor. Significant predictors included developmental delay (associated with longer diagnostic delay) and family history of similar symptoms (associated with shorter diagnostic delay). Other variables, including sex, nuclear versus mitochondrial genetic etiology, seizures, cardiomyopathy, muscle weakness, and stroke-like episodes, did not reach statistical significance.

**Supplementary Table 2**

Family Demographics of individuals who were asymptomatic but were tested because of family history. The proband was the individual who presented with symptoms and initiated cascade testing. The parent of Family 3 had an affected sibling who was not seen in our clinic. DM2=Diabetes Mellitus Type II. GDD=Global Developmental Delay, SNHL=Sensorineural Hearing Loss, N/A=Asymptomatic Individuals.

**Supplementary Table 3. Frequency and lead time of HPO terms documented prior to clinical suspicion**

Summary of all HPO terms identified in the electronic health record (EHR) with documentation before and after the point of clinical suspicion for mitochondrial disease. For each feature, the table provides the proportion of patients with documentation before and after suspicion, the number of patients contributing lead time data, and the mean and median years the feature was recorded prior to diagnosis. Inclusion in the analysis required that each individual have at least one HPO term documented before suspicion, to account for data fragmentation across records.

## Discussion

In this study of 58 individuals molecularly diagnosed with mitochondrial disease evaluated at a tertiary referral center between 2016-2025, we found that diagnostic delay remains substantial, with a mean of 8.8 years from symptom onset to diagnosis. Most of this delay (92.8% of the time) occurred before clinical suspicion was raised, while the time from suspicion to diagnosis was comparatively short. Delays have shortened across generations, with later-born individuals and those with more recent symptom onset experiencing significantly faster diagnoses. Phenotype-specific predictors also emerged: brainstem involvement and developmental delay were associated with shorter delays, whereas visual loss and later age of symptom onset were associated with longer delays. It’s unclear from the current study if this shortening is due to improve sequencing or improved clinical recognition, although we speculate both factors are contributory. Finally, analysis of Human Phenotype Ontology (HPO) terms demonstrated that diagnostically informative features, including seizures, hypotonia, developmental delay, and stroke-were frequently documented years before suspicion, underscoring a wide diagnostic window and repeated missed opportunities for recognition. ^16–18^

Diagnostic odyssesys contribute to patient morbidity, and in some cases mortality, as well as patient frustration, mental health exacerbations, and a rise in healthcare costs. In this study, five individuals from four families were diagnosed with primary mitochondrial disease through cascade testing after symptom onset in a close relative, highlighting both the utility and the ethical complexity of testing unaffected family members. ^19^ This may be further compounded by variable penetrance and variable expressivity seen across many different primary mitochondrial diseases. ^20^ Heteroplasmy levels varied widely between asymptomatic (46–92% in blood; 55% in urine) and affected relatives (39–95%), underscoring the unpredictable relationship between genotype and phenotype. For example, a child who suffered a cerebral infarction during an orthopedic procedure was found to carry the m.3243A>G MELAS variant at 77% heteroplasmy, prompting consideration of testing his asymptomatic siblings, both under age 15. While prophylactic arginine may have a limited role in modulating stroke risk, evidence for its use and efficacy is lacking; this raises questions about the clinical utility of testing asymptomatic minors, particularly when heteroplasmy complicates prognostication and a positive result may cause anxiety, stigma, or fear of discrimination despite protections such as the Genetic Information Nondiscrimination Act. ^21,22^ The five cascade cases likely underestimate the true prevalence, as some relatives decline testing. ^23^ More broadly, diagnostic timelines have improved across generations, likely driven more by the widespread adoption of exome and genome sequencing than by earlier clinical recognition. ^24^ By applying a conceptual model that partitions the diagnostic odyssey into pre- and post-suspicion intervals, we showed that most delay occurs before suspicion, when canonical features are present but unrecognized. Leveraging EHR-derived HPO terms with natural language processing further demonstrated how diagnostically informative phenotypes are systematically overlooked in real time. Taken together, these findings echo prior literature on the variability and inconsistency of diagnostic trajectories in monogenic disease, while also pointing to priority areas for intervention, ranging from ethical frameworks for cascade testing to the development of AI-driven tools that could assist recognition when suspicion is absent. ^25–27^

This study has several limitations. First, it reflects a single-center cohort from a tertiary referral clinic, which may bias toward more complex or severe cases and limit generalizability. Access to testing and awareness may differ across health systems, underscoring questions about equity and healthcare access across different socioeconomic, geographical, and demographic groups. ^28^ Second, dates of symptom onset and suspicion were abstracted from medical record to inconsistent documentation, and fragmentation across health systems. ^29^ Although we used a standardized definition of clinical suspicion, its ascertainment inevitably relied on documentation practices that may vary between clinicians and specialties. Third, while we applied NLP for systematic HPO extraction, missingness or data fragmentation in clinical notes and variation in terminology could underestimate the true prevalence or lead time of some features. Finally, the relatively modest sample size limits our ability to stratify by individual genotypes or to fully assess predictors across rarer subgroups.

Despite these limitations, this study highlights several key implications. First, improving recognition of mitochondrial disease requires tools and training to ensure that canonical features documented in the EHR, such as seizures, hypotonia, and stroke, trigger earlier suspicion. Second, our findings provide retrospective evidence for the potential of a genotype-first approach, in which sequencing is pursued reflexively when certain combinations of phenotypes are present, thereby bypassing delays in clinical recognition. This approach may be complicated by presence of variants of unknown significance, the incomplete ability of genetic sequencing alone to catch all primary mitochondrial diseases, and the challenges with mtDNA variant and heteroplasmy interpretation. Third, the successful use of NLP-based Human Phenotype Ontology extraction illustrates how automated phenotypic modeling could be deployed in real time to flag patterns highly suggestive of mitochondrial disease, prompting earlier referral and testing. ^25,30,31^ Fourth, the marked reduction in diagnostic delays across generations suggests that earlier recognition is possible and may be further accelerated through awareness campaigns, integration of phenotype-driven decision support, and expanded access to sequencing. Fifth, by constructing a large, systematically annotated but censored dataset, this study provides a foundation for the development and validation of machine learning and AI tools aimed at shortening diagnostic delay in the future. Future directions of this work may involve integrating phenotype recognition algorithms in EHR as well as targeted educational outreach across different specialties. Finally, these findings align with broader calls to standardize definitions of diagnostic delay and to apply conceptual frameworks across rare genetic diseases, thereby enabling benchmarking, cross-cohort comparisons, and evaluation of interventions designed to shorten the diagnostic odyssey.

## Conclusion

In summary, diagnostic delays in mitochondrial disease are driven primarily by missed recognition of canonical features rather than limitations in testing. By applying a standardized framework and leveraging EHR-derived phenotypes, this study provides retrospective evidence that a genotype-first strategy or automated phenotypic modeling could substantially shorten the diagnostic odyssey. Embedding these approaches into clinical practice offers a path toward earlier, more equitable diagnosis and improved care for individuals with mitochondrial disease.

## Notes

### Competing Interest Statement

The authors have declared no competing interest.

### Author Declarations

IRB 25-00728 Icahn School of Medicine at Mount Sinai Institutional Review Board

