## Supplementary material for "Drivers of Diagnostic Delay in Mitochondrial Disease: Missed Recognition of Canonical Features": Table 2 Supplement

Supplement Table 2

| **Familial Cases:** | Heteroplasmy (blood) | Presenting Symptom |
| --- | --- | --- |
| m.3243A>G Family 1: n=3 |  |  |
| Parent | 21% | DM2 |
| Sibling | 55% (urine) | N/A |
| **Child (Proband)** | **39%** | **Migraines** |
| m.3243A>G Family 2: n=3 |  |  |
| **Proband** | **77%** | **Stroke** |
| Sibling | 46% | N/A |
| Sibling | 47% | N/A |
| m.3243A>G Family 3: n=3 |  |  |
| Parent | 34% | SNHL |
| Child 1 | 60% | N/A |
| Child 2 | 77% | GDD, hypotonia, exercise intolerance |
| m.8363G>A Family 4: n=3 |  |  |
| **Parent (Proband)** | 95% | Ataxia, SNHL |
| Child 1 | 92% | N/A |
| Child 2 | 86% | N/A |
| m.14484T>C (Incidental Finding) | 100% | N/A (Tested for Congenital Arthrogryposis) |
